# A publicly available score for evaluating hospital financial standing

**DOI:** 10.1101/2020.10.31.20223750

**Authors:** Radoslav Zinoviev, Harlan Krumholz, Richard A. Ciccarone, Rick Antle, Howard Forman

**Affiliations:** Department of Internal Medicine, Yale New Haven Hospital, 20 York St, CB2041, New Haven, CT, 06510; Harold H. Hines, Jr. Professor of Medicine (Cardiology) and Professor in the Institute for Social and Policy Studies, of Investigative Medicine and of Public Health (Health Policy); Director, Center for Outcomes Research and Evaluation, Yale-New Haven Hospital; Co-Director, Robert Wood Johnson Foundation Clinical Scholars Program, Yale University; President, Merritt Research Services; William S. Beinecke Professor of Accounting, Yale School of Management; Professor of Radiology & Public Health (Health Policy); Professor in the Practice of Management; Professor of Economics (joint by courtesy); Director, YSPH Health Care Management Program; Director, MD/MBA Program; Faculty Director, Executive MBA Program (Healthcare); Yale University

**Keywords:** health economics, health policy, statistics & research methods

## Abstract

**Abstract:** *Objectives:* To create a straightforward scoring procedure based on widely available, inexpensive financial data that provides an assessment of the financial health of a hospital.

*Design:* Methodological study.

*Setting:* Multicenter study.

*Participants:* All hospitals and health systems reporting the required financial metrics in 2017 were included for a total of 1,075 participants.

*Interventions:* We examined a list of 232 hospital financial indicators and used existing models and financial literature to select 30 metrics that sufficiently describe hospital operations. In a set of hospital financial data from 2017, we used Principal Coordinate Analysis to assess collinearity among variables and eliminated redundant variables. We isolated 10 unique variables, each assigned a weight equal to the share of its coefficient in a regression onto Moody’s Credit Rating, our predefined gold standard. The sum of weighted variables is a single composite score named the Yale Hospital Financial Score (YHFS).

*Primary Outcome Measures:* Ability to reproduce both financial trends from a “gold standard” metric and known associations with non-fiscal data.

*Results:* The validity of the YHFS was evaluated by: (1) assessing its reproducibility with previously excluded data; (2) comparing it to existing models; and, (3) replicating known associations with non-fiscal data. Ten percent of the initial dataset had been reserved for validation and was not used in creating the model; the YHFS predicts 96.7% of the variation in this reserved sample, demonstrating reproducibility. The YHFS predicts 90.5% and 88.8% of the variation in Moody’s and Standard and Poor’s bond ratings, respectively, supporting its validity. As expected, larger hospitals had higher YHFS scores whereas a greater share of Medicare discharges correlated with lower YHFS scores.

*Conclusions:* We created a reliable and publicly available composite score of hospital financial stability.

**Article Summary:** Strengths and Limitations of This Study

- There is a lack of models for assessing the financial state of hospitals in a robust and systematic way using publicly available data.
- We created the Yale Hospital Financial Score, a compound financial ranking of hospitals using a diverse collection of hospital financial metrics.
- The score ranks hospitals from 0 to 100 and was validated by showing reproducibility on a pre-excluded sample and strong correlation with “gold standard” metrics
- This score has been developed to aid health policy researchers and has not yet been validated in studies of longitudinal financial outcomes

## C. Introduction

The rapidly changing American healthcare environment exerts continuous financial pressure on hospitals. There is evidence that this pressure affects patient care: studies have shown both a higher rate of adverse patient events in hospitals under financial distress^1^ and improved quality of care when profitability increases^2^. New policies, such as the introduction of quality-based reimbursement, may further propagate these adverse effects by placing additional selective pressure on already struggling institutions. Conversely, providing financial assistance to financially disadvantaged hospitals may prevent adverse events and improve care.

Empirical research on the relationship between hospitals’ financial conditions and patient outcomes is needed to inform any decision to change reimbursement policy and to evaluate any changes that are implemented. Such research faces an immediate problem in how to define and assess a hospital’s financial condition. Most modern studies rely on financial ratios, the Altman Z Score and credit ratings to assess hospital finances.

Financial ratios are readily available and are therefore most frequently used by hospital managers^3^, health policy researchers^2,4,5^, and departments of public health^6^ to assess hospital financial condition and performance. There are, however, hundreds of candidate financial ratios, and there is no generally agreed on subset on which to focus. This problem is not unique to hospitals. Financial analysts typically wade through a sea of ratios, choosing the ones that make intuitive sense to them or that have been dictated by company policy.

There has been an effort to identity meaningful composites: single scores comprised of several financial ratios. The most extensively validated publicly available composite to indicate financial distress is the Altman Z Score^7^. Altman focused on predicting the two-year potential for bankruptcy and used a statistical approach on a sample of publicly held manufacturing firms to develop the Altman Z Score.

This score, a composite of five financial ratios, achieved 90% accuracy^8^ and several modern studies have shown its continued validity^9–11^. Caution should be applied, however, in applying the Altman Z Score to assess the overall financial condition of hospitals. Revisiting the model in 2002, Edward Altman himself reminds us that the Z Score’s intended use is assessing *distress* in *manufacturing companies*^12^. Studies extrapolating its use to healthcare^5,13^ or in predicting financial success^14^ have been limited and inconclusive.

Credit rating agencies, such as Standard & Poor’s and Moody’s, also create composite scores that attempt to capture the likelihood that a firm will pay back its creditors. These agencies use a combination of statistical studies, experience and judgment to derive a rating from underlying financial statistics. Credit ratings are, to our knowledge, the best available indicator of financial health, but they are proprietary, often costly to obtain, and are simply unavailable for many hospitals. For hospitals with municipal credits, Merritt Research Services LLC produces a ranking that is both specific to hospitals and aimed at financial condition more broadly than are the credit ratings.

There is no perfect model for assessing overall hospital financial standing, and current “gold standard” composite scores are unavailable to most health policy researchers. In this paper we have undertaken a pragmatic approach to creating a simple, one-dimensional score, the Yale Hospital Financial Score (YHFS), that can be used to indicate the financial condition of a hospital. The YHFS involves a straightforward calculation that relies on widely available data that can be obtained at little or no cost. We assess the validity of the score by its ability to predict debt ratings, when they exist. We find that the YHFS explains 88.8%, 90.5% and 95.0% of the variation in three different credit ratings.

## D. Methods

Hospital financial data for fiscal year 2017 was downloaded from the publicly available Centers for Medicare and Medicaid Services (CMS) Hospital Cost Reports (HCR) database^15^. Hospital demographic data was downloaded from the CMS Hospital Compare database^16^. All healthcare institutions reporting complete fiscal information in 2017 were included in the sample. The final sample of 928 institutions was comprised of acute care hospitals, critical access hospitals, hospital districts and health systems. We partnered with Merritt Research Services LLC, an independent research and data provider, to investigate the creation of a new hospital financial model. Merritt Chief Executive Officer Richard Ciccarone provided guidance in identifying key variables in hospital financial statements and in the use of statistical models to select the variables that are most important in predicting hospital financial performance.

We studied the financial analysis literature and identified 232 financial metrics that describe all aspects of hospital operations. To select metrics that provide a complete and diverse representation of hospital operations, we examined reports on financial management, consulted financial modelling literature, reviewed financial associations with patient outcomes^2^, and inspected the methodology used by credit rating agencies^17,18^. We selected 30 ratios **(Figure 1)** that, based on our research and the criteria below, appear to be essential in describing the overall financial standing of a hospital:

**Figure 1:**
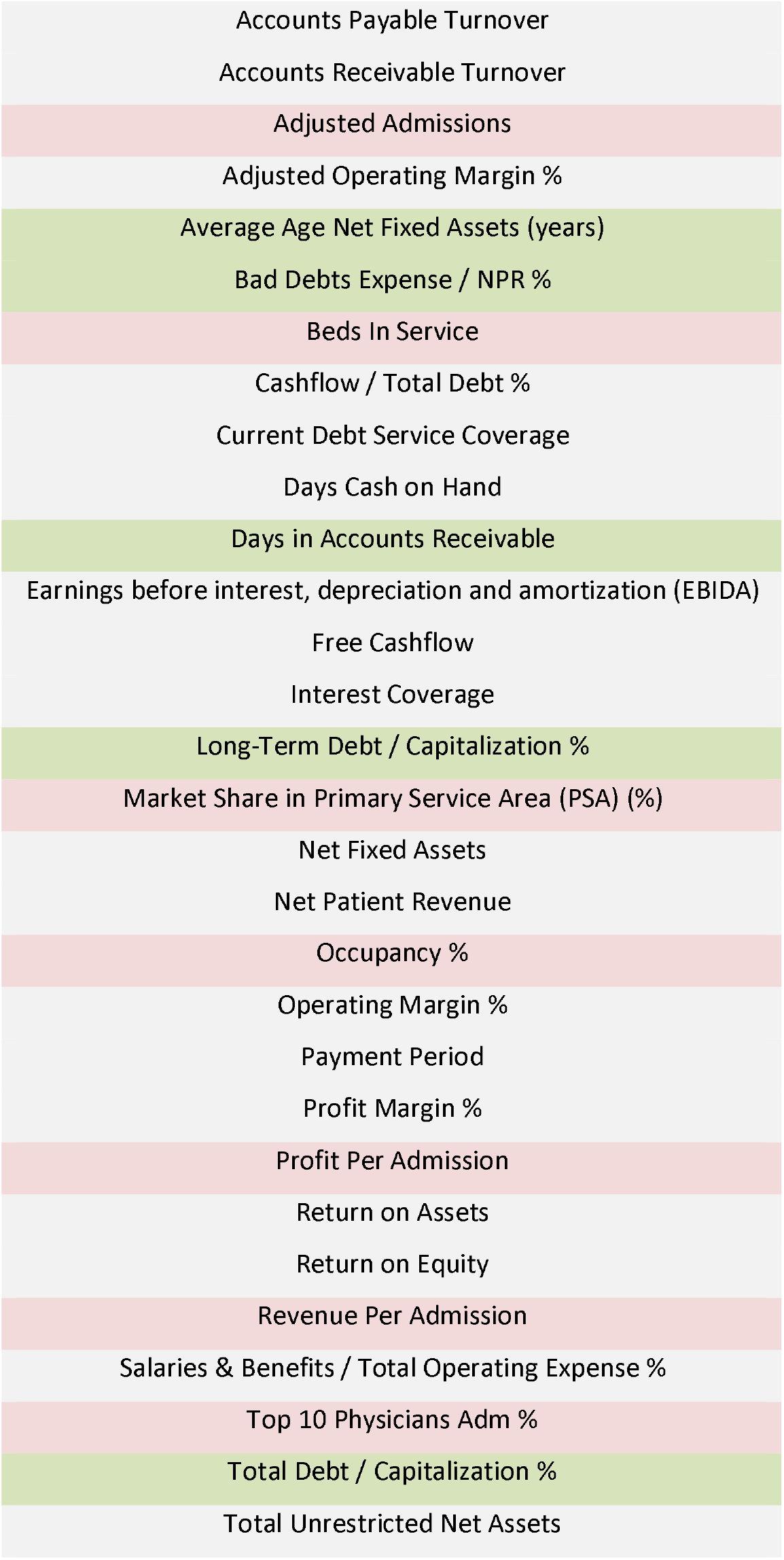
Predictors of Economic Performance. List of 30 metrics identified as strong predictors of economic performance of hospitals for use in a ranking score of hospital financial standing after initial evaluation of 232 potential candidates. Metrics in red were removed due to poor reporting rate (<10%) in the CMS Cost Reports database. Direction of metrics in green was changed to warrant positive correlation with utility.

a. Common use in bond ratings of financial institutions
b. Ability to indicate the makeup of a hospital’s patient pool (ex. Medicare/Medicaid-predominant vs private insurance)
c. Predictability of asset management in the hospital
d. Profitability, both from patient admissions and overall profit margins

These 30 ratios were computed from numeric values found in the HCR dataset using financial ratio standard equations determined by the US Generally Accepted Accounting Principles (GAAP)^19^. For example, profit margin is the quotient of net income / total revenue. Seven of the pre-selected 30 ratios could not be computed due to lack of data in the HCR database (defined as data missing for >10% of hospitals, shown in red, **Figure 1**). The directionality of 5 “negative” financial metrics was changed (shown in green, **Figure 1**) to ensure that increasing utility was positive for all variables. After the 23 ratios were computed, 90% of the 1,075 hospitals in the sample were randomly selected as the analysis group and the remaining 10% were excluded from the analysis for later use in verification.

We used Principal Coordinate Analysis (PCoA) to examine trends in the dataset. Values were standardized and converted to natural numbers (≥0, µ=0, σ=1). PRIMER analytic software (Vers. 6. Lutton, UK: PRIMER-E Ltd, 2009) was then used to graph the relationship between component measures and calculate the Euclidian distances between pairs of variables **(Figure 2)**. The Euclidean distance threshold required to form variable groups was 50. Variables that clustered together **(Figure 2)** were thought to be redundant. To eliminate redundant variables, we used Principal Components Analysis (PCA). PCA of the standardized dataset returned 9 Principal Components with a minimum Eigenvalue of 1 that describe nearly 80% of the variation in the data. For each variable cluster found through PCoA, variables that were weighed more heavily in the PCA were retained and the others were removed from the analysis. Variables that formed unique, single clusters in the PCoA were automatically selected for inclusion. In total, ten variables were selected to include in the hospital ranking score **(Figure 3)**.

**Figure 2:**
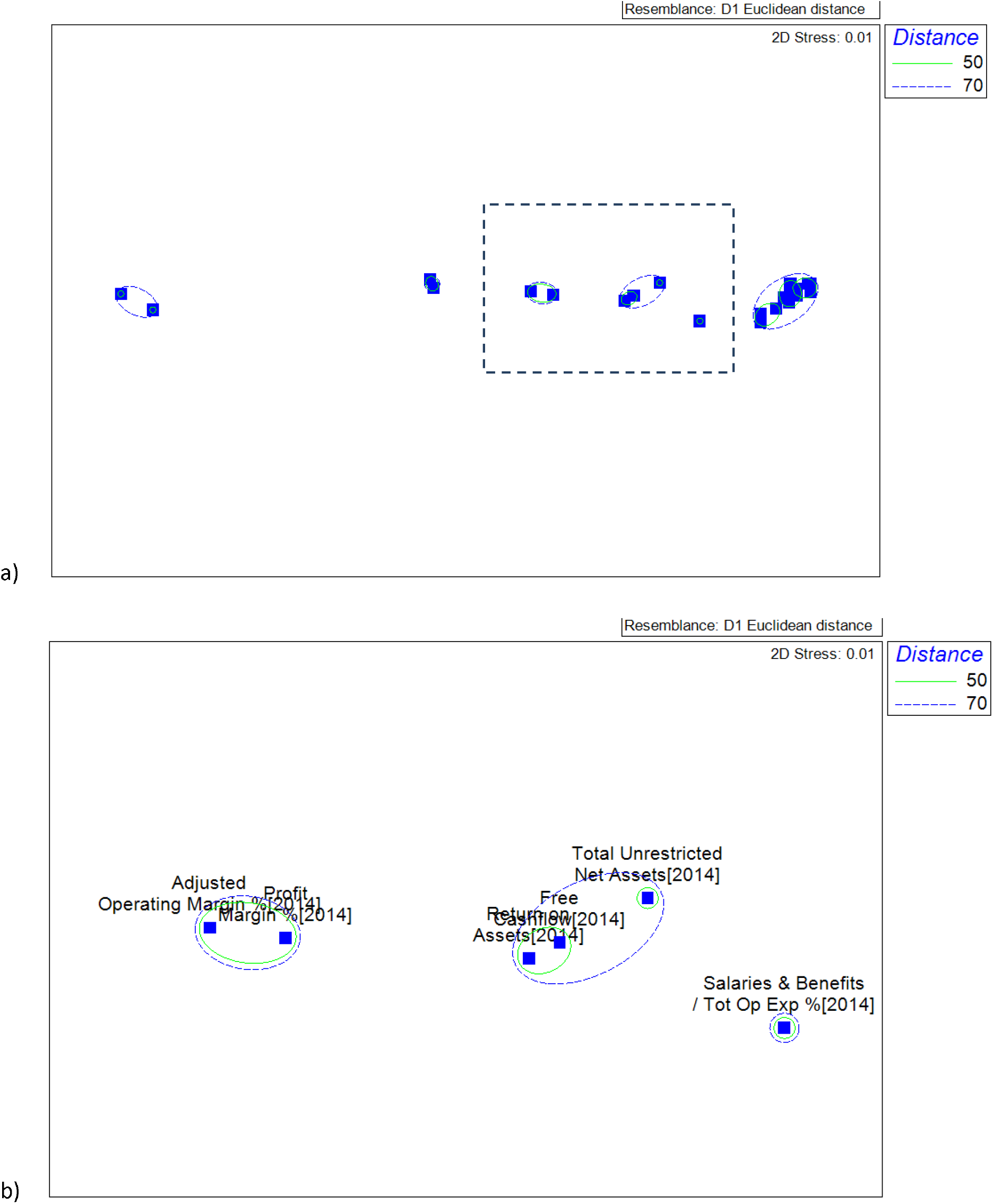
Principal Coordinate Analysis (PCoA) of hospital financial metrics. (a) Cluster analysis of the Euclidean distances; data point labels are removed for visual clarity. (b) Magnified section of the mid-right sector of the cluster analysis with data point labels added.

**Figure 3:**
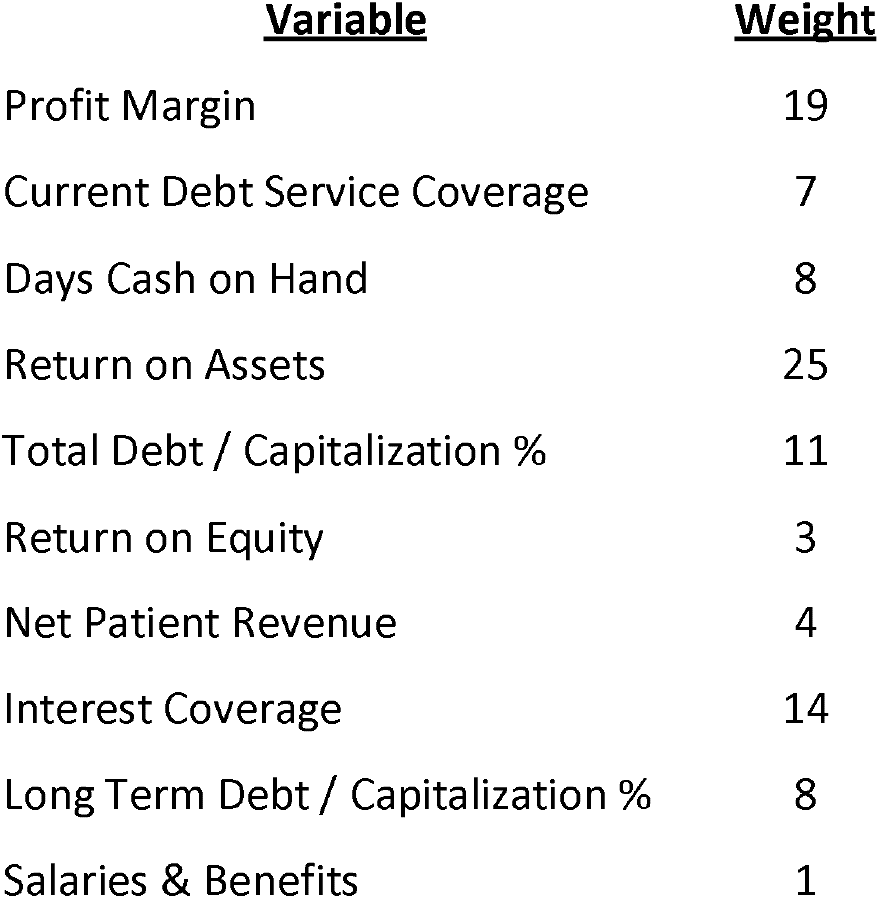
The Yale Hospital Financial Score. The ten financial ratios comprising the YHFS were derived from hospital audit reports, with the corresponding weights used in calculating the composite model.

Moody’s Credit Rating is the most widely used credit rating for hospitals and was therefore selected as the presumed gold standard for our analysis. As such, we used Moody’s Credit Rating to determine the relative weight for each variable. We ran an ordinary least squares (OLS) regression model of the numeric Moody’s Credit Rating on the 10 variables and used the coefficient for each variable as the relative weight of that variable in the model. The weights were then standardized to a total of 100 as shown in **Figure 3**. Values were ranked within a variable using the Excel *percentrank* function and the final financial score was calculated as the sum product of ranked values using the weights determined from the analysis.

Patient and public involvement in the study was facilitated by open discussion in the medical and business communities. In seeking to create a novel means of assessing hospital financial standing, the authors solicited open feedback regarding the study’s design and the choice of metrics. During the first round of attrition, 30 metrics were selected from a list of 232 with input from financial experts in the Yale School of Management. Subsequent variable selection through statistical analysis was inspired by work from biostatisticians with whom the study was discussed. Open discussions regarding the design of the YHFS ultimately led to the recruitment of Mr. Richard Ciccarone and Prof. Rick Antle from the private and academic financial sectors.

Evaluation of the model was conducted using OLS regression analysis. All statistical analyses in this study were performed using STATA statistical data analysis software (Vers. 15. College Station, TX: StataCorp LP, 2013).

## E. Results

We constructed a compound score for evaluating hospital financial stability as outlined in the methods, henceforth referred to as the **Yale Hospital Financial Score (YHFS)**. We assessed the validity of this model through several tests. We had reserved 10% of the hospitals in the original dataset for validation. After computation of the YHFS, an OLS regression model of the YHFS on the ten financial variables for the reserved hospitals predicts 96.7% (r^2^=0.967) of the variation in our model. We then compared our model to existing proprietary credit scores, used as gold standards. The CreditScope Rank is a proprietary rating constructed by Merritt Research Services LLC, based on their evaluation of hospital finances. There was a significant correlation between our model and the CreditScope Rank **(Figure 4)** and OLS regression showed that the YHFS explains 95.0% (r^2^=0.950) of the variation of the CreditScope Rank. The YHFS predicted 90.5% (r^2^=0.905) of the variation in Moody’s and 88.8% (r^2^=0.888) of the variation in S&P’s bond ratings.

**Figure 4:**
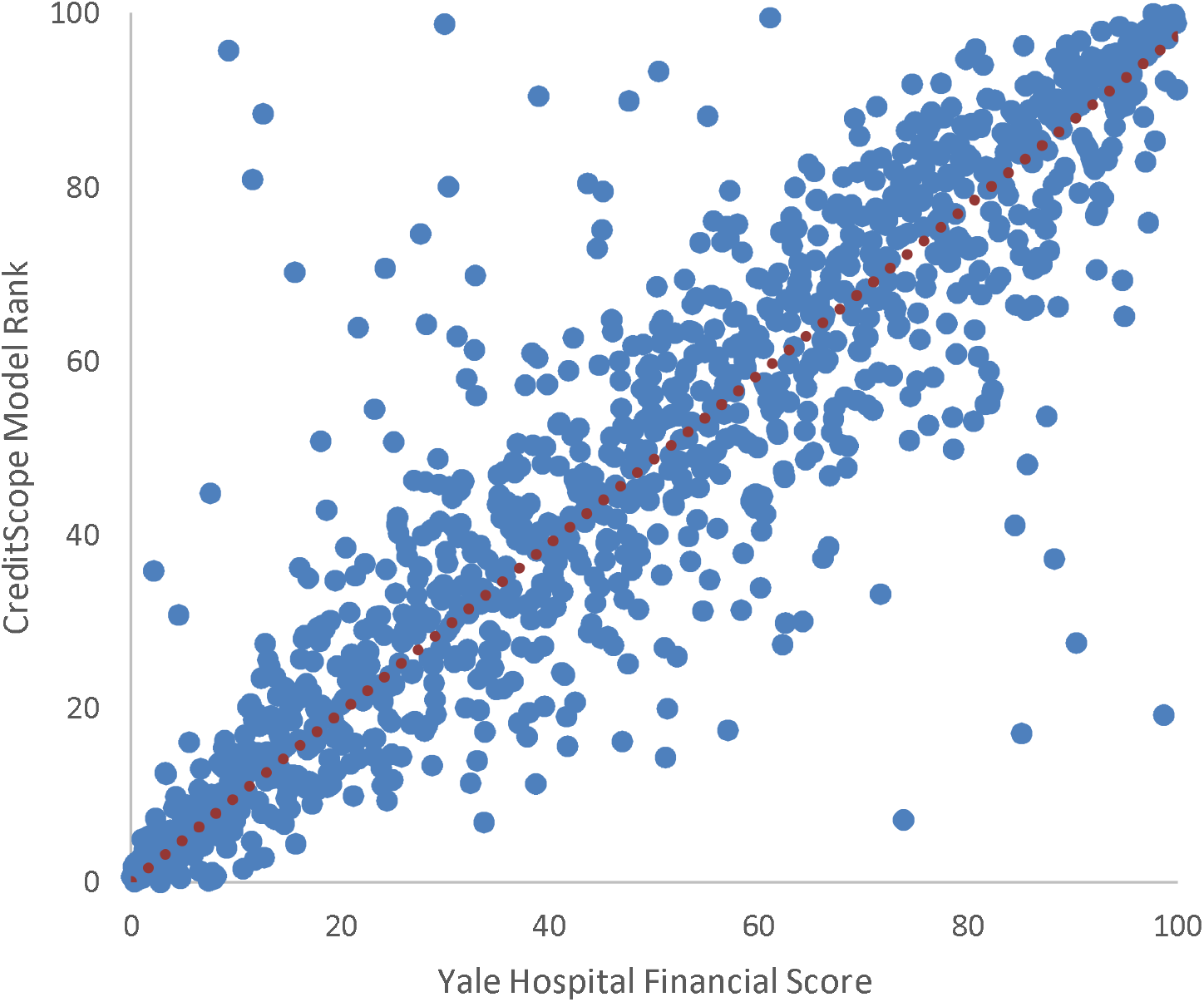
Validating the Yale Hospital Financial Score. Graphical comparison of the Yale Hospital Financial Score to the CreditScope Model for financial standing of hospitals.

To further test the validity of our model, we extend the analysis beyond fiscal data, using a multivariate OLS regression to examine the effect of non-monetary factors on the financial stability of a hospital. We found that larger hospitals are at lower risk of default, with a 1.7-point higher YHFS score for every additional 100 beds (0.017, 95% CI 0.01 to 0.02, p<0.001). Conversely, the share of Medicare patients seen at a hospital is associated with a 0.4-point lower YHFS for each additional percent of Medicare patients as a share of total annual discharges (−0.36, 95% CI −0.49 to −0.22, p<0.001). We found no association between hospital financial standing and teaching status of the hospital (6.61, 95% CI −4.19 to 17.4, p=0.23).

## F. Discussion

There is currently no perfect model for evaluating the financial standing of hospitals. We undertook a systematic approach in creating a publicly available and transparent financial score. Starting with a list of 232 financial metrics pertaining to all aspects of hospital finance, we selected the 30 most pertinent ratios and used statistical analysis to exclude redundant variables. This method identified 10 financial metrics that were used to construct a single score for a hospital’s risk of default compared to other hospitals. After these 10 metrics were isolated, we evaluated each of the ten ratios included in our model for their indispensability in predicting a hospital’s risk for default. *Return on assets* is a strong indicator of a hospital’s ability to generate revenue while limiting its costs, a reflection of payment models in the value-based-care era. Analysis of hospital data has suggested that high *profit margin* can signify fewer competitors and a favorable payer mix^20^. High *return on equity* allows a hospital to finance growth whereas low return on equity can make it susceptible to liquidation or acquisition^21^, an increasingly common practice in the healthcare industry^22^. Not surprisingly, measures of revenue and profit represented a total 51% of the YHFS score. Next to revenue and profit, debt is the second most important indicator for a hospital’s risk of default. Debt is a complex variable because more established and successful companies can take on higher debt at lower rates that can be leveraged to increase revenues in excess of expenses. Complicating the matter further, hospitals are at higher risk of bad debt than other operating entities because they are obligated to provide services before compensation can be guaranteed. To incorporate the intricate impact of debt on risk of default, we included four separate measures of debt: current debt service coverage, interest coverage, *total debt* and *long-term debt to capitalization*, which collectively represent 40% of the YHFS score. Unlike other businesses, hospital may not collect payment until months or even years after a service is rendered due to complexities with insurance reimbursement. Therefore, sufficient *cash on hand* is critical for hospitals, who cannot rely on immediate or totally predictable payment, yet too much can signify poor management of funds^23^. Finally, hospitals compete for a highly skilled workforce, and the *salaries and benefits* paid by a hospital reflect its ability to generate capital and attract talent, in turn drawing in patients and improving outcomes.

After constructing the model, we used three accepted means of evaluation to show its validity^24^: (1) by extrapolation to a portion of the initial dataset that is excluded from the model’s construction, (2) by showing a high correlation to a gold standard, and (3) by replicating known associations with non-fiscal data. Since there is no agreed-upon gold standard in this field, we used accepted and widely used credit ratios. We first tested the association between the YHFS and raw financial data. We found that this dataset predicts 96.7% of the model’s variation in a small sample of hospitals excluded from its creation, suggesting that our methodology is generalizable. Although there is no gold standard for the scoring of hospital financial standing, credit scores predict financial instability and are sometimes used to model financial success. We found that our model is highly correlated with credit scores created by CreditScope (95.0%), Moody’s (90.5%) and S&P (88.8%). In the absence of a gold standard, these results support the value of our model.

We then studied Moody’s model more closely to assess where our model differed. To assess the impact of differences over time, we computed the YHFS for hospitals using 2014 data and compared it to Moody’s Credit Ratings for those same hospitals in 2014, then examined how the most varied hospitals have fared over this 5-year period. We found that the average difference between the score predicted by our model and the numeric Moody score in 2014 was 18/100 points (σ=15). This was uniformly distributed across each Moody’s score decile. A primary difference between the two models is that Moody’s focuses on absolute revenue whereas we used metrics that evaluate a hospital’s ability to generate and utilize funds. Secondly, we believe that the management and repayment of debt is paramount in the healthcare industry and used a more inclusive assessment of debt by including several measures of debt-management. Finally, we included measures of asset utilization and efficiency that have been shown to be important in avoiding bankruptcy in the healthcare industry. While our score tended to closely resemble the score assigned by Moody’s, the score for 12 hospitals differed by more than 50/100 points between the two scales. In all 12 cases the Moody’s score was >50 points *higher* than the score predicted by our model. Since 2014, Moody’s has downgraded the credit ratings for 6 of these 12 hospitals and withdrew its rating for 1 hospital. In 2 of the 6 cases, the credit rating was downgraded by several tiers and in a 3^rd^ case a hospital was downgraded from a top tier credit rating to its second lowest rating. One hospital was sold, one merged with another health system and a third laid off 42 employees. In the hospitals whose credit scores differed by more than 25/100 points between the two models, only 7 hospitals were scored higher by our model. For 4 of these 7 hospitals, Moody’s has significantly increased their credit rating since 2014 and for the remaining 3 hospitals, Moody’s credit rating has not changed.

Our third test of validity was using our model to replicate known associations between hospital finances and non-fiscal data. Studies have shown that larger hospitals tend to be more efficient than smaller ones^25^ because of economies of scale, better recognition, or other factors. A 13-year longitudinal study of financial distress in public hospitals published in 2014 found an increased likelihood of financial distress in hospitals where Medicare payments comprised a greater share of total revenues^26^. Finally, the finances of a hospital have not been found to be associated with the hospital’s teaching status^27^. All three of these findings were replicated by our model score. Hospital scores in our model are higher for larger hospitals, lower for hospitals with a larger share of Medicare patients and are not associated with the teaching status of a hospital.

## G. Implications

Numerous studies have shown an association between hospital financial distress and worse patient outcomes^1,2^. A model of hospital finances would be invaluable in health policy research, though there is currently no perfect model. Existing methods are often proprietary and are either created with limited information or extrapolated from non-healthcare industries. An ideal model should draw data from diverse financial domains and have proven validity, rely on publicly available data, be easy to construct and provide a single composite score to facilitate comparison to other hospitals and to patient outcomes data. In this study we have used publicly available data to create a financial score using transparent, systematic methodology that may be easily reproduced and used in health policy research. We openly acknowledge the limitations of the YHFS: current “gold standards” assess risk of default and there is no perfect way in certifying the validity of a new model. We have used currently available methods and associations to create a putative score for hospital financial standing. Such a model may screen for hospitals at risk for default, provide information regarding the fiscal effect of new policies, suggest correlations between financial performance and patient outcomes and indicate how different revenue streams affect overall performance.

## H. Conclusion

We outlined a method for constructing a composite score for assessing hospital financial standing using publicly available data. We used three approaches to validate this composite, which we called the YHFS. First, we tested its reproducibility by showing a high degree of association between our model and financial data for hospitals excluded from the sample set used in its construction. Second, we tested its accuracy by showing a high degree of association between this model and proprietary credit ratings that are the current presumed gold standard. Finally, we showed that this model can predict variation in non-monetary data known to be associated with hospital finances. We believe that the YHFS may be useful for assessing the financial standing of healthcare institutions and we hope that this publicly available score may aid health services researchers in future evaluations of hospital finances.

## Data Availability

All data used in this study was obtained from publicly available sources as detailed in the methods section.

## K. Author Contributions

Primary data analysis and preparation of the manuscript were executed by Radoslav Zinoviev, MD, MBA (corresponding author). Dr. Howard Forman, MD, MBA, was the primary advisor for this work. Dr. Harlan Krumholz, MD, and Prof. Rick Antle, PhD, were reviewing authors. Mr. Richard A. Ciccarone, MPA, assisted with data analysis and metric design.

## L. Acknowledgements

The authors would like to express their gratitude to the faculty of the Yale School of Management for the helpful discussions in designing this study, and to Dr. Naveen Venayak and Dr. Maggie Stoeva for their invaluable guidance on statistical analysis.

